# Clinical and Genetic Spectrum of Fanconi Anaemia in Australia and New Zealand

**DOI:** 10.1101/2024.10.21.24315893

**Authors:** Hannah Fluhler, Elissah Granger, Michael Sharp, Caitlin Harris, Mark Mckinley, Sarbjit Riyat, Christine Krieg, Andrew Deans, Chris Fraser, Siobhan Cross, Tina Carter, Lisa Worgan, Adam Nelson, Lucy C. Fox, Jillian Nicholl, Alison Attwood, Lorna McLeman, David Hughes, Rachel Conyers, Omar Kujan, Eunike Velleuer-Carlberg, Adayapalam Nandini, Wayne Crismani

## Abstract

Fanconi anaemia (FA) is a rare genetic condition which predisposes to progressive bone marrow failure, a specific spectrum of malignancies including head and neck squamous cell carcinoma (HNSCC), and an array of other clinical manifestations. The clinical and genetic spectrum of FA in Australia and New Zealand remains relatively undescribed in the literature and is limited to case reports. In this study, we conducted the first comprehensive investigation of FA within this region combining cohort data and case reports, aiming to elucidate its diagnostic patterns, clinical manifestations, and genetic characteristics. Our findings reveal a positive correlation between national testing rates and case detection, across states and territories of Australia, suggesting that targeted testing strategies may enhance the identification of FA cases. Further, our analysis demonstrates that the physical and genetic profiles of FA patients in Australia and New Zealand resemble those observed in other international cohort studies. This study emphasises the continued significance of cytogenetic testing for FA while stressing the need for heightened awareness among medical professionals. FA, once perceived primarily as a paediatric condition, now demands vigilance across all age groups due to advances in medical care and complementary detection methods such as high-throughput sequencing. This study also underscores the heightened susceptibility of individuals with FA in Australia and New Zealand to HNSCC, consistent with observations in other regions worldwide.

## Introduction

Fanconi anaemia is a rare genetic condition caused by mutations in a common DNA repair pathway. FA predisposes affected individuals to a spectrum of disorders such as progressive bone marrow failure, early onset cancer, endocrine issues, physical abnormalities and reduced fertility (L. Frohnmayer, 2021; Tsui & Crismani, 2019). The traditional physical phenotypes which can be associated with FA include <u>v</u>ertebral anomalies, <u>a</u>nal atresia, <u>c</u>ardiac anomalies, tracheoesophageal fistula, o<u>e</u>sophageal atresia, renal structural anomalies, limb anomalies – and <u>h</u>ydrocephalus (VACTERL-H association) and abnormal <u>p</u>igmentation including café au lait spots, <u>h</u>eight abnormalities, <u>e</u>ndocrine disorders, <u>n</u>eurological disorders, <u>o</u>ral lesions, and <u>s</u>hort stature (PHENOS) (Fiesco-Roa et al., 2019). The clinical presentation of these associations is heterogeneous between individuals, FA siblings with the same pathogenic variants, and even identical twins, which can often make diagnosis difficult. Three or more of the 8 VACTERL-H associations only occur in 12% of FA people. Four or more of the six PHENOS associations occur in 9% of FA people (Fiesco-Roa et al., 2019). In a large literature review of FA cases, 21% of people do not have an abnormal phenotype (Fiesco-Roa et al., 2019), which can lead to cases going undetected. Therefore, cases of FA which are not diagnosed in early childhood due to an absence of overt malformations would have insufficient surveillance for life-threatening diseases to which they are predisposed such as bone marrow failure and specific malignancies such as acute myeloid leukaemia or head and neck squamous cell carcinoma (Alter et al., 2018; Altintas et al., 2022; L. Frohnmayer, 2021; Kutler et al., 2003; Niraj et al., 2019; Rosenberg et al., 2003; Shimamura & Alter, 2010).

The median age of diagnosis for FA is 6.7 years, based on data from 1,497 case reports (Shimamura & Alter, 2010) mainly due to progressive bone marrow failure. However, the diagnosis of FA may be earlier for more severe cases. Diagnostic testing for FA uses a chromosome breakage assay, which remains the gold standard functional test (Auerbach, 2009, 2015; Auerbach & Wolman, 1976; Bogliolo et al., 2019; D. Frohnmayer et al., 2014; Kimble et al., 2018; Pinto et al., 2009). The chromosome breakage assay performed with the patient’s cells is a useful diagnostic tool as it provides a functional assessment of genomic instability – the underlying molecular defect in FA – in the presence of specific DNA crosslinking agents, diepoxybutane (DEB) or mitomycin C (MMC). Another functional test is based on the finding, that FA cells arrest in the G2/M phase of the cell cycle due to inter-strand crosslinks (Heinrich et al., 1998; Knies et al., 2017). The assay for G2/M phase arrest may be easier to implement than the chromosome breakage test however it is not unique for FA. Therefore, the combination of the clinical suspicion and a positive chromosome breakage test remains mandatory to make the diagnosis FA. Genetic testing can be used to identify the gene containing the FA-causing variants. However, a challenge with diagnostics assays that use peripheral blood as the source of cells or DNA, is the issue of somatic mosaicism which occurs in 15-25% of patients (Castella, Pujol, Callén, Ramírez, et al., 2011; Gregory et al., 2001; Gross et al., 2002; Hamanoue et al., 2006; Kwee et al., 1983; Lo Ten Foe et al., 1997; Mankad et al., 2006; Nicoletti et al., 2020; Pinto et al., 2009; Soulier et al., 2005; Timmers et al., 2001), because any spontaneous mutation or recombination events that restore the function of the mutated gene in a haematopoietic stem cell results in that cell having a proliferative advantage, and becoming the dominant genotype in the peripheral blood. The diagnostic challenge associated with somatic mosaicism may be overcome by skin biopsy and fibroblasts assay for diagnostic tests. A skin fibroblast biopsy is routinely taken for the Zero Childhood Cancer Program (ZERO2) testing for whole genome sequencing as part of the inherited bone marrow failure and related disorders (IBMDx) study, so all BMF patients can have germline testing via this method once they have consented.

When a positive functional test is obtained, it is prudent to identify the genetic cause of FA because it is complementary evidence supporting the diagnosis. Further, pathogenic variant identification may assist with: i) testing of potential related stem cell donors and family planning (L. Frohnmayer, 2021; Verlinsky, 2001; Zierhut et al., 2014); ii) predicting the course of the disease in an individual, surveillance for specific malignancies(Alter et al., 2007). The identification of a pathogenic variants that cause FA may also be useful to determine eligibility for gene therapy trials and new therapeutic trials.

FA is caused by the loss of function of any one of the known 23 *FANC* genes, including *BRCA1/FANCS* and *BRCA2/FANCD1* (Frohnmayer, 2021; Niraj et al., 2019). FA is an autosomal recessive disease except for *FANCB*, which is X-linked (Meetei et al., 2004), and *FANCR/RAD51*, for which there are rare dominant subtypes (Ameziane et al., 2015). The *FANC* genes act in a common genetic pathway, sometimes referred to as the FA/BRCA pathway, which are known traditionally for their role in inter-strand crosslink repair (Knipscheer et al., 2009) but also has established roles in other types of DNA repair including fork protection, R-loop metabolism and meiosis (Crismani et al., 2012; Kolinjivadi et al., 2020; Schwab et al., 2015). This pathophysiology makes patients with FA exquisitely sensitive to agents that cause inter-strand crosslinks in DNA such as platinum-based chemotherapies. This FA/BRCA DNA repair pathway is involved in the repair of double strand breaks *via* homologous recombination. The FA/BRCA DNA repair pathway is also involved in the repair of DNA double strand breaks (DSBs) *via* homologous recombination (Nakanishi et al., 2005; Richardson et al., 2017). This FA/BRCA pathway role in the repair of DSBs may also explain why radiotherapy can cause negative effects in FA patients, as ionising radiation can cause DSBs (Alter, 2002; Birkeland, 2011; L. Frohnmayer, 2021; Tan et al., 2011).

Carrier rates for FA vary between populations but are estimated to range from 1/67 to 1/181 (Castella, Pujol, Callén, Trujillo, et al., 2011; D. Frohnmayer et al., 2014; Rosenberg et al., 2011). No publications are yet to provide the prevalence of FA in the Australian and New Zealand populations. These populations are composed of a unique mixture of people from Australian Aboriginal and Torres Strait Islander groups, Maori and Pacific Islands peoples, European ancestry and countries throughout Asia, among other regions of the world. This paper presents, for the first time, the clinical spectrum of FA cases from Australia and New Zealand and provides an estimate of FA cases in Australia from national testing data.

## Results

### Testing results in Australia and prevalence

National testing data from 927 chromosome breakage tests was aggregated from three laboratories. There were 843 tests performed over the period from 2010-2023 in a laboratory in Queensland, 61 tests from 2018-2024 from a laboratory in South Australia and 23 tests performed in 2023 from a laboratory in Western Australia. Positive tests occurred at a rate of 6.1% (n = 57/927), while 7.9% (n = 73/927) were inconclusive (Fig. 1). In the 2021 Australian Census, 3.2% of the population identified as Aboriginal (2.92%), Torres Strait Islander (0.13%) or both (0.14%). In the testing data, where ethnicity information was available, we found that Aboriginal people represented 6.2% (21/341), Torres Strait Islanders represented 0.59% (2/341), and people who identified as both Aboriginal and Torres Strait Islander represented 1.2% (4/341). Non-indigenous people represented 92.1% (314/341).

**Figure 1.**
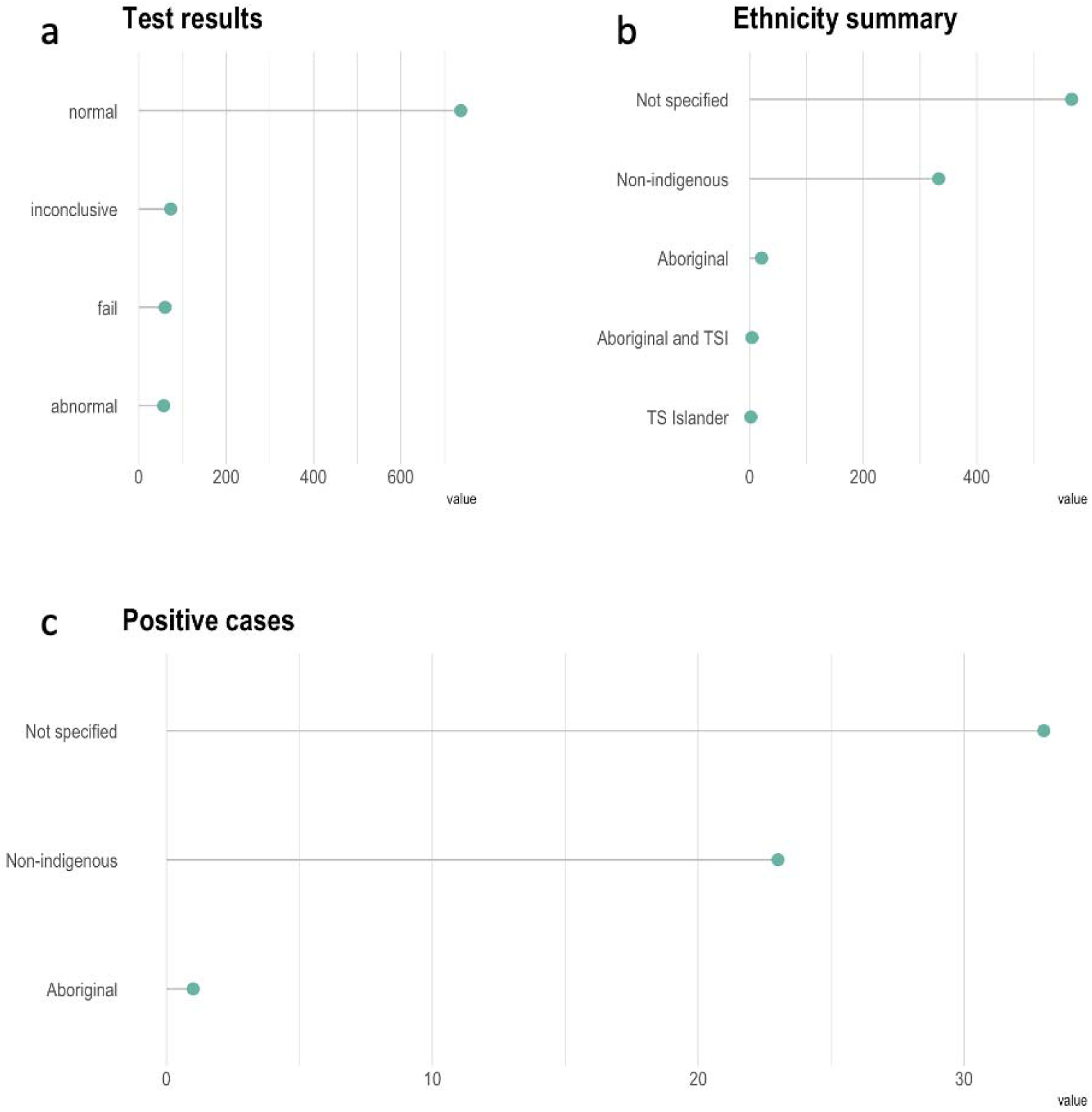
Chromosome breakage testing results. Chromosome breakage test results are summarised. a) Most test results are normal or ‘negative’ for FA. b) Tests report sometimes contain information about the ethnicity of the people tested. Testing of first nations people appears to be proportional to their fraction of the Australian population based on census data. c) Most abnormal or ‘positive’ cases for FA have no information about their ancestry, followed by those that are non-indigenous.

The median age at which individuals tested positive was 7 years (Fig. 2). Using this data, we attempted to estimate the prevalence of FA in Australia. With several conservative assumptions, we extrapolated the number of cases of FA in Australia to be approximately 189 or a frequency of 1 in 141,169 people (see Supplementary Text for calculations and assumptions). For the people where information about biological sex was available, 377 were female and 446 were male and 104 tests did not have this information. The rate of testing of females and males appeared to be different (X-squared = 5.8, df = 1, p-value = 0.016), showing that females are tested less frequently than males (see Discussion). However, no significant difference was detected in the number of positive cases (males n = 27, females n = 21, NA = 9, X-squared = 0.75, df = 1, p-value = 0.39).

**Figure 2.**
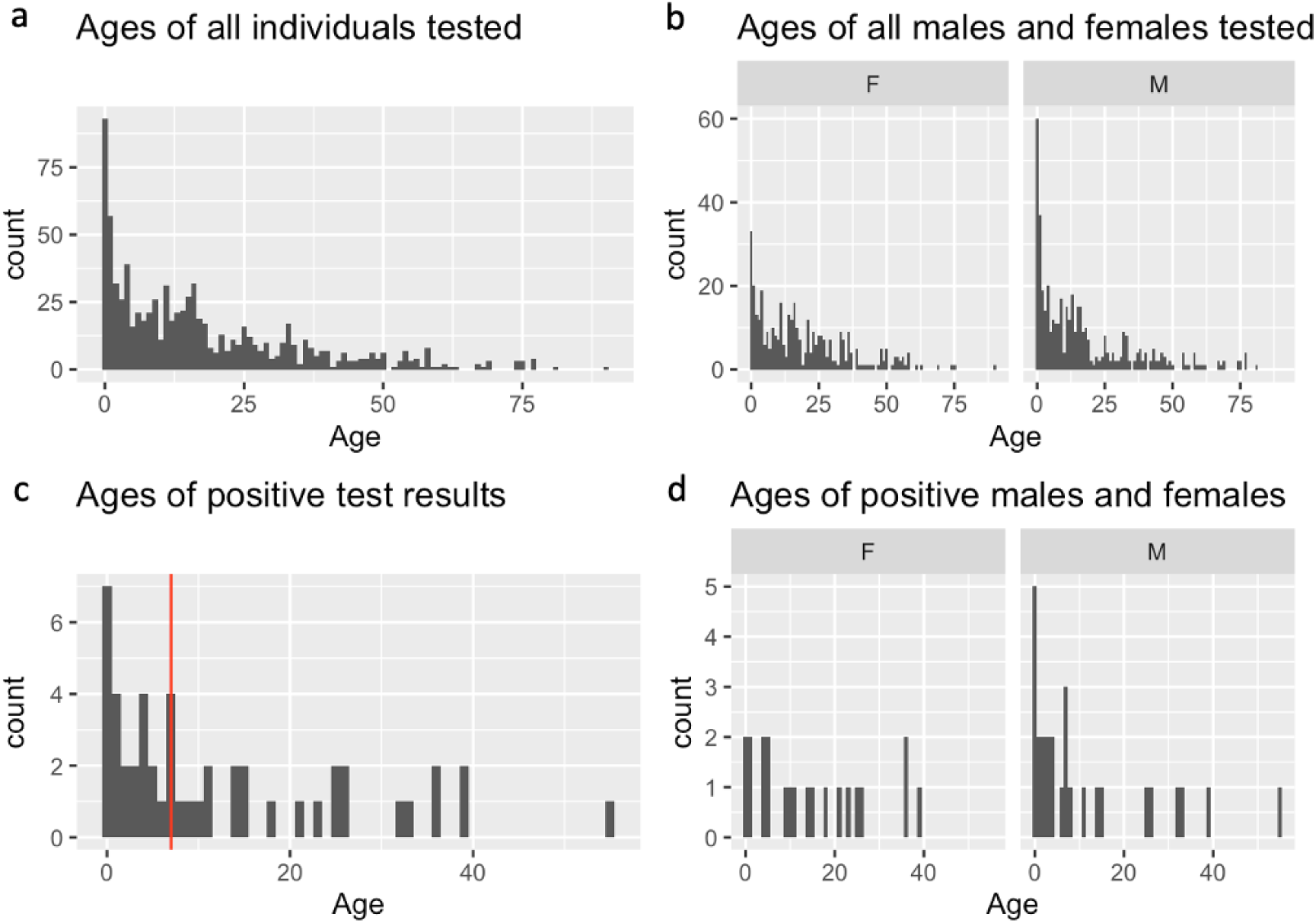
Testing ages for Fanconi anaemia in Australia. a) All tests and the ages of the individuals, where available (n = 831), are presented as a histogram. b) The same tests are separated as a function of biological sex. c) The ages of all individuals who returns a positive test presented as a histogram with the median represented by a vertical red line. d) The same positive tests are presented separating results into the biological sexes of the individuals as either male or female. There is a male who tests positive at age 55. Details of this individual with a homozygous mutation in *FANCJ* is discussed below and previously published (Stevens et al., 2016).

The highest rate of testing per million people came from the state of Queensland (Fig. 3). Notably, this is the state where the laboratory is located (see Discussion). The highest rate of positive tests per million people in the population also came from Queensland. We noticed that increased testing in a state or territory tends to positively correlate with more cases of FA being detected in Australia, with no particular indication that the highest test rates in this dataset have reached saturation. Testing rates were noticed to increase over time in the testing data from the Queensland laboratory. Specifically in the year 2017, there appeared to be an increase in testing numbers which was sustained in subsequent years. From 2010-2016 the average number of tests was 28 ± 17, and from 2017-2023 the average number of tests was 82 ± 23, which represents a 2.9-fold increase in tests per year. In the corresponding periods the number of positive tests was 2.7 ± 1.4 (2010-2016) and 4.1 ± 1.7 (2017-2023) (see Discussion).

**Figure 3.**
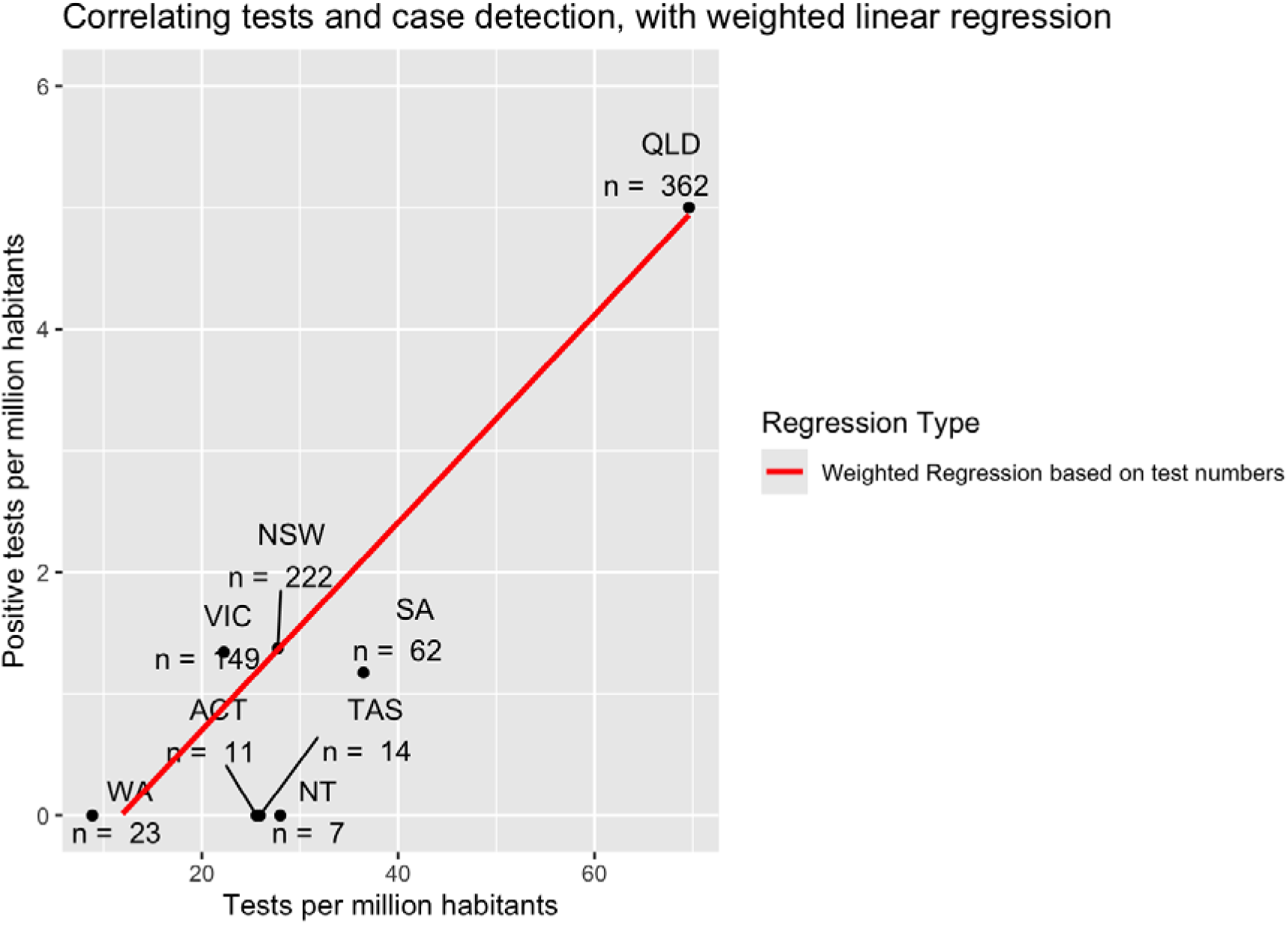
Increased testing rates positively correlate with detection of FA cases. More testing for FA in the states and territories of Australia positively correlates with more cases being detected without signs of saturation, based on weighted linear regression (red line) using total test numbers per state. Note that tests for WA were only from one year (2023) from the WA laboratory, tests from SA were from six years (2018-2024), and tests from other states and territories are from the Qld laboratory over 2010-2023.

### Demographics of individuals with FA in Australia and New Zealand

A total of 12 individuals with FA from Australia and New Zealand provided informed consent to share information. The median age of diagnosis in this special cohort was only 1 year of age (n=12). Enriching this data with the identified published Australian and New Zealand cases (n=8) the median shifts to 4.5 years of age (n=20; 8 females, 12 males). These median ages of diagnosis are unexpectedly younger than what is observed in the national testing data, which highlights a potential sampling bias with respect to who participates in research (see Discussion). Physical manifestations were skewed towards the upper limbs, short stature, and kidneys (Figure 4). These physical findings are relatively consistent with other studies (Altintas et al., 2022; Fiesco-Roa et al., 2019).

**Figure 4.**
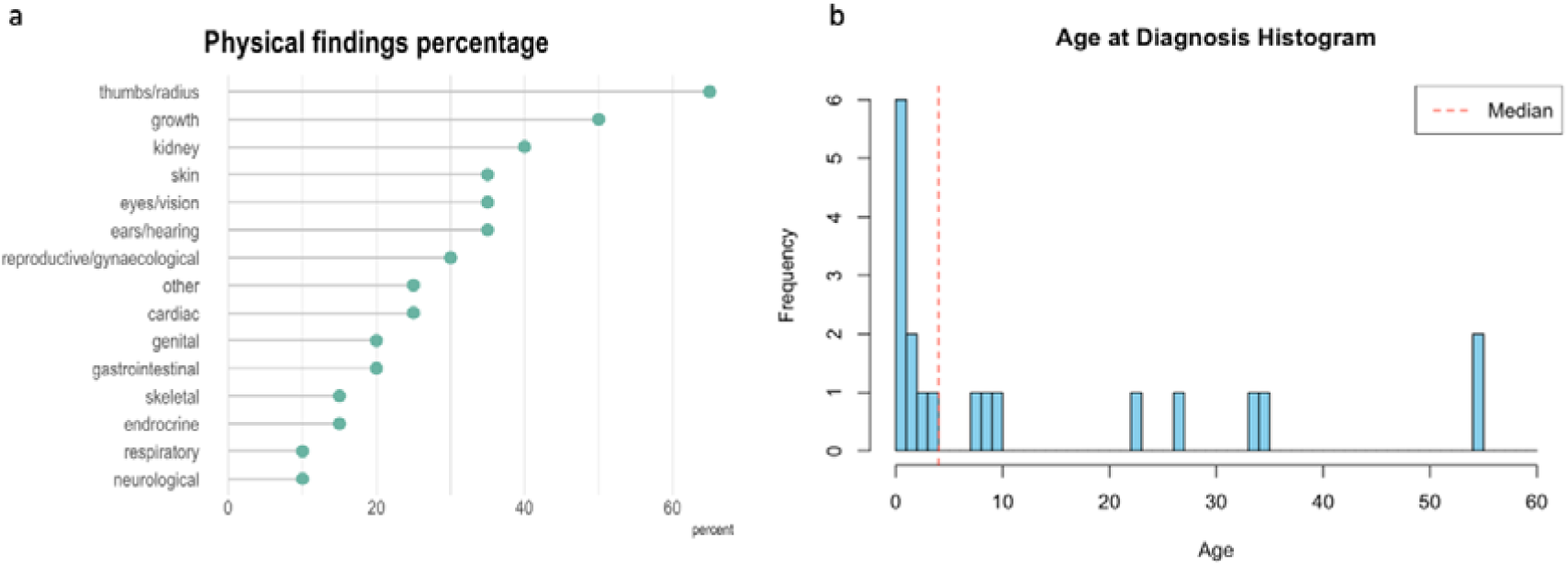
Physical manifestations of FA are dominated by upper limb and renal issues as well as short stature. a) Physical manifestations of FA in are consistent with previous studies. Issues of the upper limbs are the most common physical feature of FA seen in most people in this cohort and the literature, followed by short stature in half of the people. Renal issues are also common. b) Histogram of the age of diagnosis of the combined cohort and cases from the literature.

**Table 1.** Characteristics of the people. Aggregated information is presented about the characteristics of the participants in this study. “N” refers to the number of participants for whom information was available for a particular characteristic; in some instances, data could not be obtained for all fields for all participants. “Value” refers to how many people present a given characteristic from those for whom information was available. Percentage is (N / Value) * 100.

| Characteristic | N | Value | Percentage | Median age | Cohort |
| --- | --- | --- | --- | --- | --- |
| Positive cases, median age at FA testing of cohort and literature | 20 | - | - | 4.5 | Study cohort and literature |
| Positive cases, median age at FA testing of cohort | 12 | - | - | 1.5 | Study cohort |
| Positive cases, median age at FA testing of literature | 8 | - | - | 34 | Literature |
| Positive age, median age at FA testing from pathology laborato | 770 | - | - | 7 | National testing data |
| Biological sex - male | 20 | 8 | 40% |  | Study cohort and literature |
| Biological sex - female | 20 | 12 | 60% |  | Study cohort and literature |
| Age at onset of haematological manifestations (0-3yo) | 12 | 3 | 25% | - | Study cohort |
| Age at onset of haematological manifestations (4-10yo) | 12 | 5 | 42% | - | Study cohort |
| Age at onset of haematological manifestations (>10yo) | 12 | 1 | 8% | - | Study cohort |
| Percentage transplanted | 11 | 5 | 45% | - | Study cohort |
| Median age at bone marrow transplant | 5 | - | - | 7 | Study cohort |
| No malformations | 19 | 2 | 11% | - | Study cohort and literature |
| 1-2 malformations | 19 | 3 | 16% | - | Study cohort and literature |
| 3+ malformations | 19 | 14 | 74% | - | Study cohort and literature |

### Time to onset of bone marrow failure, haematopoietic stem cell transplant and cancer

A total of 9/12 people had experienced decreased blood counts, which most frequently presented between 4-10 years of age (Table 3). A total of 5/11 people had a bone marrow transplant. Regarding cancer, a total of 6/20 people had a malignancy. The cumulative incidence of solid tumours by the age of 40 was 20% (4/20), including individuals under 40, some of whom may not have reached the age at which the event could manifest, thus reflecting both observed cases and censored data. The most common solid malignancy was head and neck squamous cell carcinoma (HNSCC) which occurred in 3/20 of the cases, all of which were in young adults. The increased risk of HNSCC is consistent with the risk seen in international cohorts with adult FA people (Alter et al., 2018; L. Frohnmayer, 2021; Kutler et al., 2003, 2016; Velleuer et al., 2020; Velleuer & Dietrich, 2014). 4/12 people self-reported having persistent oral lesions. There were two cases of bowel cancer, including gastrointestinal adenocarcinoma (Ip et al., 2021). There was a separate case of gastric tubular adenoma (Ip et al., 2021). There were two cases of hepatocellular carcinoma (Blombery et al., 2021; Stevens et al., 2016). 1 person had a melanoma. One person had T-acute lymphoblastic leukaemia (Ryland et al., 2020). One person had metastatic head and neck squamous cell carcinoma which was excised with surgery and radiation.

Australia and New Zealand’s populations are composed of substantial diversity with a blend that is unique based on its geography and pre- and post-colonial timelines. According to the Australian Bureau of statistics 2021 Census data, ancestries in Australia are: Oceanian, which includes Australian Aboriginal, Torres Strait Islander, Australian, New Zealand and pacific islands; North-West Europe, Southern and Eastern European, North African and Middle Eastern, South-East Asian, North-East Asian, Southern and Central Asian, Peoples of the Americas, Sub-Saharan African (*Australian Bureau of Statistics*, 2022). Participants ancestries were skewed towards Caucasian (Figure 5), which is consistent with the non-indigenous status of most individuals in the diagnostic testing data (Figure 1). Two individuals in our study had aboriginal ancestry, and this study is the first that reports on individuals with FA of Australian aboriginal ancestry. In New Zealand, according to the official data agency, Stats NZ Tatauranga Aotearoa, the major ethnic groups are European, Maori, Pacific Peoples, Asian, MELAA (Middle Eastern/Latin American/African) and other ethnicity. No individuals recruited into this study reported Maori heritage. However, a patient with Maori ethnicity was identified but had died of Acute Myeloid Leukaemia prior to the study (personal communication, Siobhan Cross).

**Figure 5.**
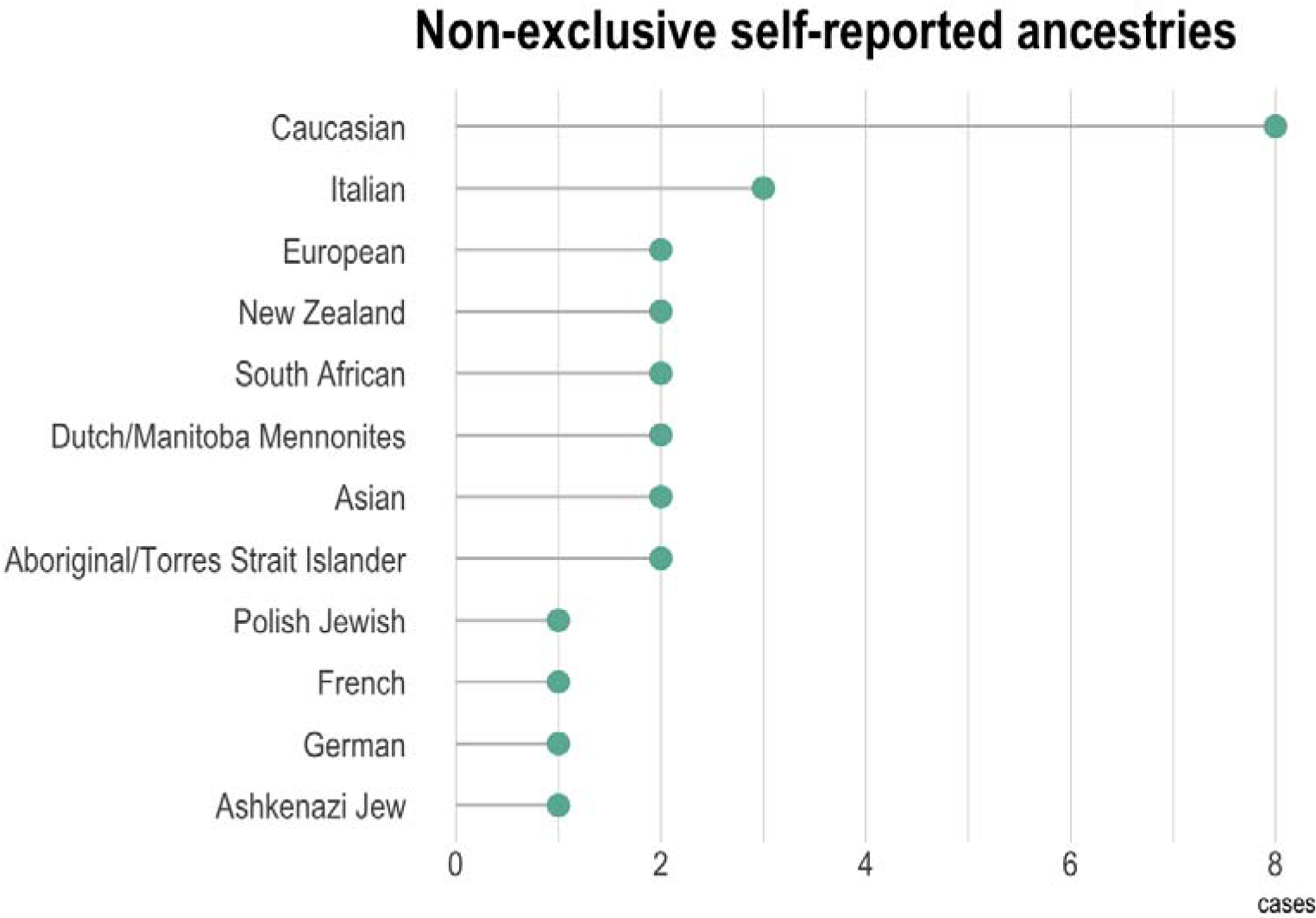
Ancestral backgrounds of study participants are predominantly Caucasian. Self-reported ancestries of the cases and literature (n = 15). Individuals can report more than one ancestry.

### Genetic subtypes of individuals with FA in Australia and New Zealand

The genes with pathogenic variants were skewed towards *FANCA* (7/14 resolved cases). The second most common gene was *FANCD2* (3/14), and other rare subtypes made up the rest (Figure 6). The people with mutations in *FANCA* and *FANCD2* do not appear to be related based on the *FANC* gene variants and should not account for the over representation of either of these genes. The only exception was a single case of FA attributable to homozygous variants in *FANCA*. The published twins with homozygous mutations in *FANCJ* were reported to be from non-consanguineous parents. We identified one novel variant, *FANCD2* (c.1691_1692del), which would cause a premature stop codon at Ser564 in the translated protein (Figure 7).

**Figure 6.**
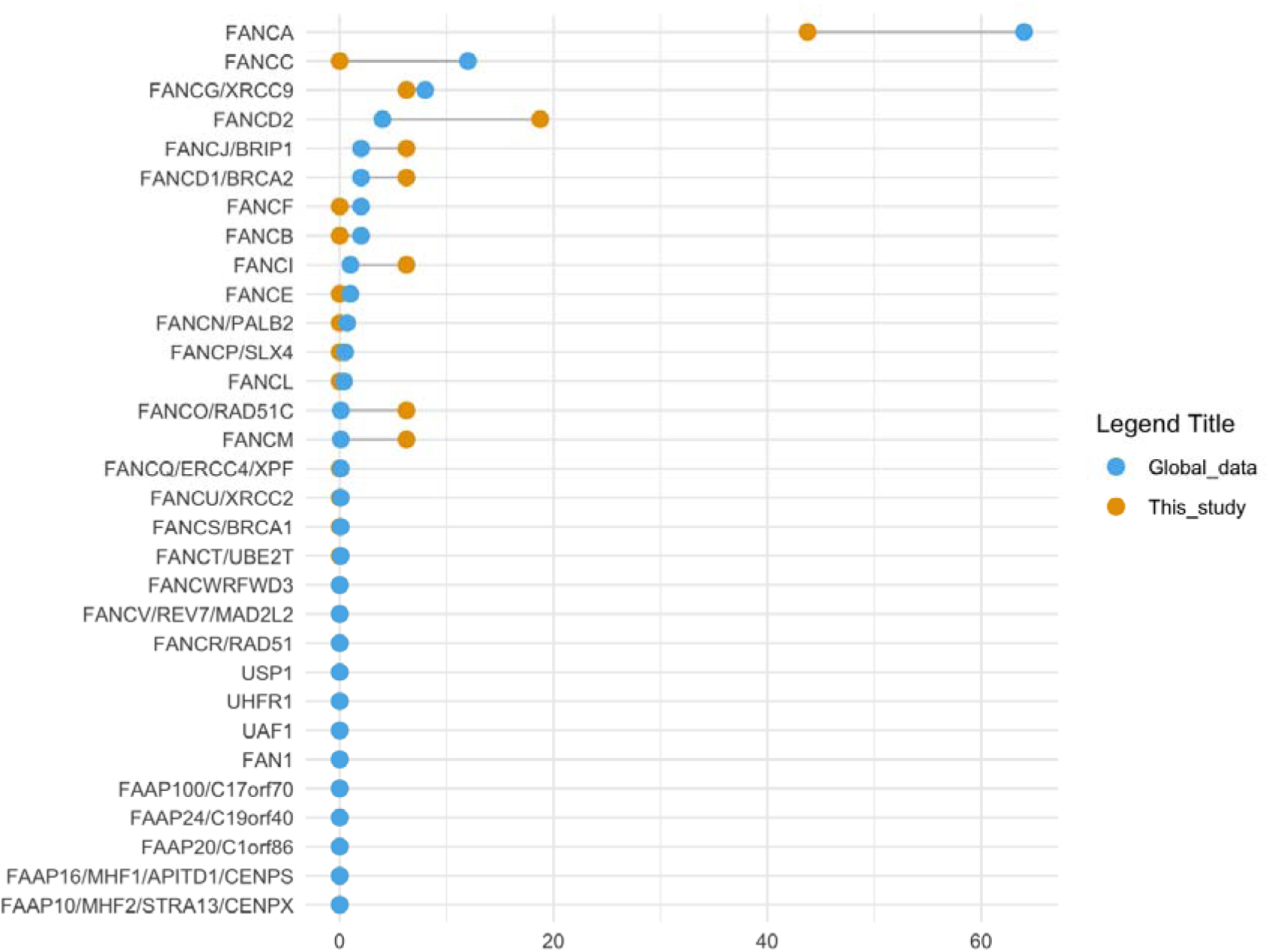
The most frequently mutated gene in FA cases is *FANCA* consistent with other cohorts. Gene mutation frequencies from cases of FA are plotted in descending frequencies. Observed frequencies from the 20 Australian and New Zealand cases are similar to the global data from Frohnmayer et al.

**Table 2.**
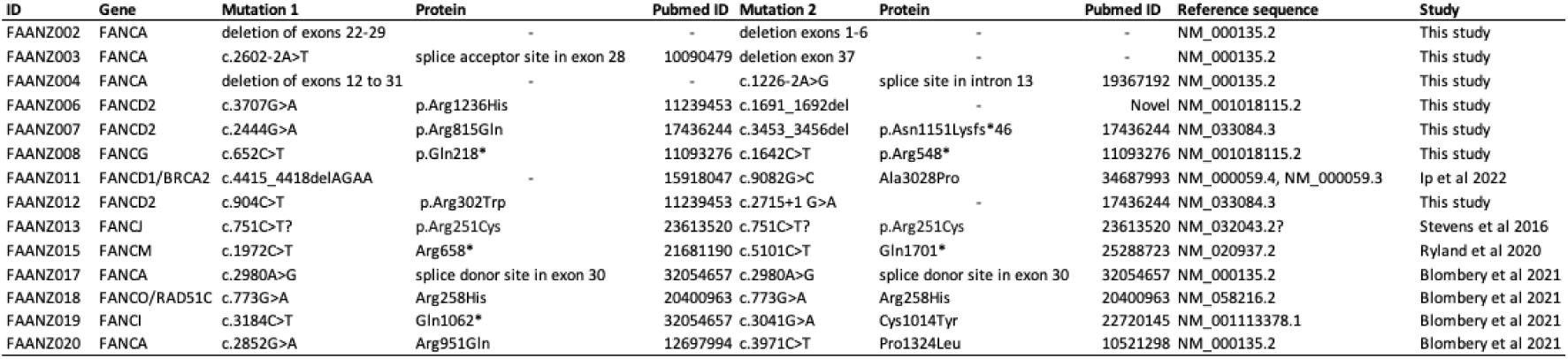
Genetic variants in *FANC* genes in Australian and New Zealand people with Fanconi anaemia.

**Figure 7.**
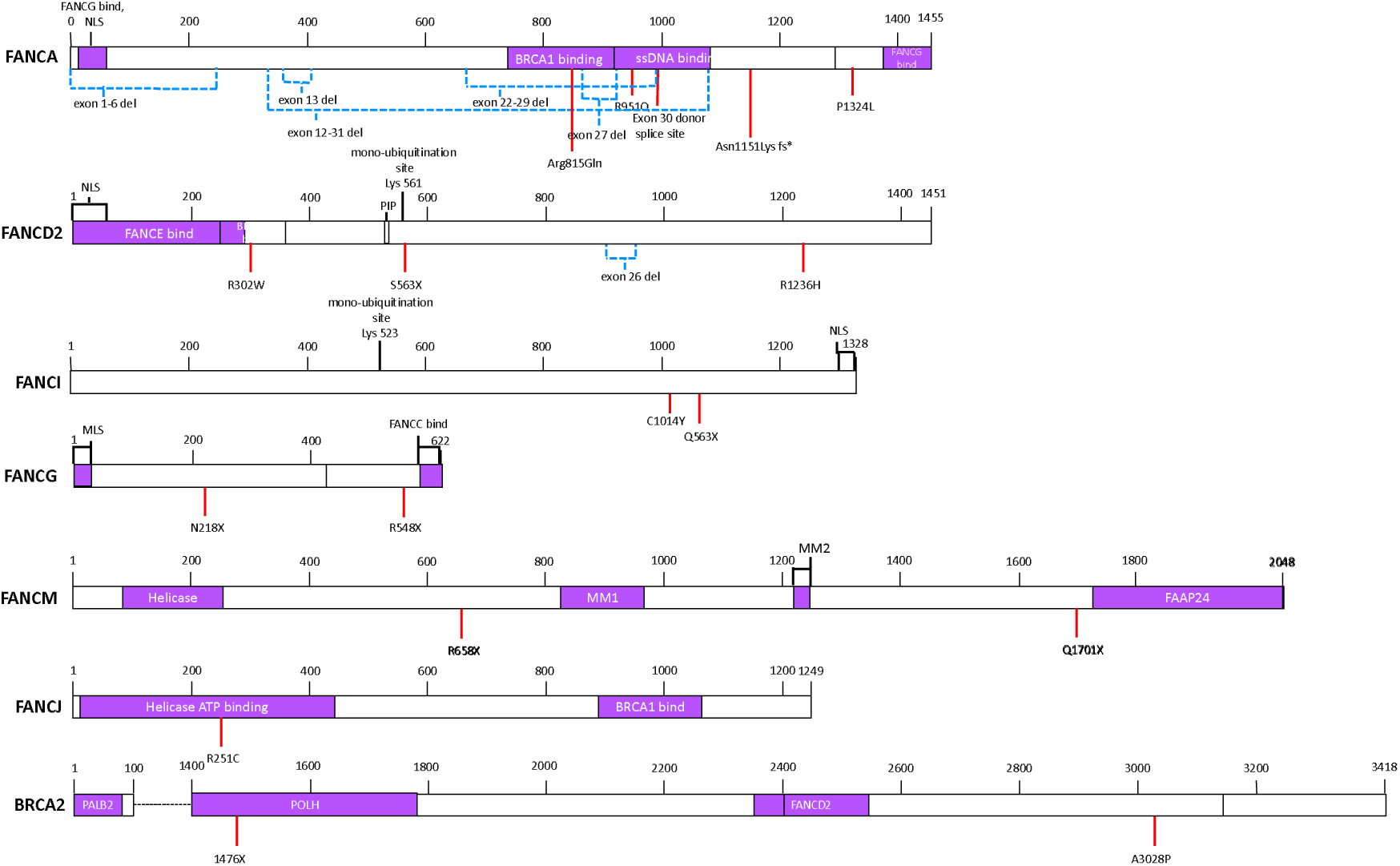
Mapping of genetic variants onto the proteins. Genetic variants are shown on each of the respective proteins.

## Discussion

### Age of diagnosis

This study describes the first cohort of people with FA from Australia and New Zealand and substantial testing data from Australia. The main findings are that diagnostic rates of FA in most of Australia appear to be similar to that of the US, when considering the median age of diagnosis, which is 7.

#### How does the diagnosis of FA in Australia and/or New Zealand compare to other countries?

The testing data from the cytogenetic lab suggests that with a median age of diagnosis of 7, Australia is similar to the US (Shimamura & Alter, 2010). Nevertheless, higher testing rates in across states and territories of Australia positively correlate with more cases being detected per million people. This is consistent with the hypothesis that more cases of FA will be detected if there is more testing for FA where a higher index of suspicion may be reasonable, and guidelines from the Fanconi Cancer Foundation provide a useful resource to consider which patients could be tested for FA (Supplementary Fig. 1) (L. Frohnmayer, 2021). The highest testing rates in Australia, and the highest number of cases detected per capita was in Queensland. It is unclear what drivers this increased testing. One speculative possibility could be due to referral systems which is in some way tied to the physical proximity to the testing laboratory. Another non-exclusive speculative possibility may be a group of doctors who elect to test for FA more frequently due to a particular awareness of the condition.

We observed that the number of tests for FA increased from 2017 onwards. Interestingly this approximately 3-fold increase in tests performed correlated with a 53% increase in cases detected per year. One possible reason for this large increase in testing numbers is because these tests are requested by bone marrow transplant teams who are systematically testing all patients with aplastic anaemia for FA prior to a bone marrow transplant as part of clinical protocols and followed updated guidelines (Killick et al., 2016) suggesting testing for FA prior to transplant in adults with inherited aplastic anaemia. Therefore, this highly prudent and sensible test, would only identify a small number of previously undetected cases of FA. The value however of those who it does identify as having FA is extremely high as it means that these individuals will receive the appropriate conditioning regimen for their transplant, and appropriate follow up and care post-transplant.

We have provided the first estimate of Australia’s rate of FA cases to be 1/141,169. This is relatively similar to some historical estimates from other countries. However, our estimation is conservative for the following reasons: i) There are more inconclusive tests than abnormal/positive in the data. A portion of these tests may reasonably be cases where hypomorphic mutations in *FANC* genes have led reduced function of the FA pathway in these individuals, predisposing them to pathologies of FA; ii) We do not know if all tests from the states and territories discussed in this study go to these Australian testing laboratories or to international laboratories. iii) Cases of FA will be missed as roughly 25% of people have no traditional phenotypes and receive a diagnosis due to health issues later in life such as head and neck cancer, haematological manifestations, or infertility.

### Possible sex bias in testing

It is possible that males are tested at higher rates than females, particularly in their first year of life. It is unclear what might cause this and is unlikely to be occur through haematologists as it is rare to have cytopaenias in infancy. One possibility is that male children with growth faltering can be more likely to be referred for evaluation (Grimberg et al., 2011). One speculative contributing factor could be that because the genitourinary system is sometimes affected in FA (L. Frohnmayer, 2021), when a clinician detects such an abnormality, a test for FA may be requested. And therefore, because differences in sex development are easier to detect in someone who is genetically male (46XY), because the genitalia are external and visible without sophisticated imaging methods that might be required for females, this might lead to more tests for FA in newborn males, particularly if low set thumbs are also present.

### Genetics of FA in Australia and New Zealand

The distribution of genetic subtypes, *i.e.* which genes are mutated, is similar to what has been seen in other studies. *FANCA* is the most common subtype. *FANCD2* was the second most frequently occurring subtype, which has been observed in a cohort of 201 Spanish FA patients (Bogliolo et al., 2019). However, with the small number of samples, it is unclear if this would be maintained as the cohort increases in size prospectively as *FANCC* is the second most common subtype in a US-based cohort (Auerbach, 2009). Notably, the three cases from our cohort with pathogenic *FANCD2* variants are not related.

### Cancer in the FAANZ cohort

Head and neck cancer is of particular concern to people with FA. Oral lesions have been reported by four individuals and three young adults have been diagnosed with HNSCC. This is exceptionally higher than the lifetime risk of the general population. Most of the cancers described here, are from the cases added from literature searches. The cancer cases from the literature skews towards less common genetic subtypes of (*FANCD1/BRCA2*, *FANCJ*, *FANCM*) and are published cases where the genetic diagnosis was not made until the person had experienced chemotherapy-induced toxicity. It might have been reasonable to expect myeloid malignancies in this cohort, based on international literature, however this was not the case.

### Early diagnosis and barriers to access health care system

Individuals with FA have 500-800-fold higher than general population in developing HNSCC (Kutler et al., 2003). Therefore, the current guidelines suggest that routine surveillance should begin at the age of 10, which is the earliest age reported for HNSCC diagnosis in the literature and should be coordinated with specialised oral pathologists (L. Frohnmayer, 2021). FA patients often have numerous oral lesions, and it requires a healthcare provider with extensive experience in evaluating and managing HNSCC to distinguish between suspicious and non-cancerous oral lesions in these patients. Oral clinicians, such as those with backgrounds in oral medicine, oral surgery, and head and neck surgery, may serve as frontline healthcare professionals in the detection and monitoring of oral lesions. This has been recognised as oral manifestations can be early indicators of the condition in the subset of patients who pass through childhood without detection of their condition, and who have hypomorphic variants (L. Frohnmayer, 2021; Lach et al., 2020). In addition, oral brush biopsy-based cytology has been found to be a reliable adjunct in the surveillance of patients diagnosed with oral potentially malignant disorders (Idrees et al., 2022), including those with FA (Velleuer et al., 2020).

### Limitations of the study

It is not possible to establish how many tests might have been sent to laboratories overseas.

## Conclusion

We have described here the first cohort of individuals with FA in Australia and New Zealand, including nationwide FA testing data from Australia. We also describe FA testing data. Our findings suggest that the detection rate of FA increases with more testing, which is of huge clinical importance for individuals who would otherwise be missed and therefore not receive appropriate care. We also found that the presentation of FA in Australia and New Zealand is particularly similar to other countries when considering features such as physical features, genetic subgroups, and an increased rate of bone marrow failure and HNSCC compared to the general population. Therefore, as Australia and New Zealand do not have their own clinical guidelines for the diagnosis and management of FA, but clinical features are similar to what is already described, it would be reasonable to use the international Fanconi Anemia Clinical Care Guidelines.

## Data Availability

All data produced in the present work are contained in the manuscript

## Acknowledgements

We wish to thank all study participants and their families who were involved in this study. We also thank all members of the Crismani laboratory for their comments on the manuscript. W.C. receives a fellowship and funding related to this work from the Australian National Health and Medical Research Council and Victorian Cancer Agency (MCRF21006, GNT1129757, GNT2030115). SVI receives Operational Infrastructure Support from the Victorian State Government. We thank the Fanconi Cancer Foundation for permission to use of their infographic about FA.

## Methods

### Ethics, participant recruitment and clinical records

This study was approved by the Royal Children’s Hospital Human Research Ethics Committee (2019-282). Study participants provided written informed consent. We recruited 12 participants with an FA diagnosis from Australia and New Zealand. All published FA cases from Australia and New Zealand were also compiled (Blombery et al., 2021; Ip et al., 2021; Merriman et al., 2002; Ryland et al., 2020; Stevens et al., 2016). Consenting study participants completed questionnaires and access to medical records. Individual chromosome breakage test results and pathogenic *FANC* gene variant information was obtained where possible. Phenotypic anomalies were considered absent if not reported. Details of any statistical tests that were used are described in the text in context.

### Chromosome breakage testing

We obtained chromosome breakage test data for the periods 2010 to 2023 from Queensland Health. We aimed to estimate how many cases of FA might be in Australia. We reviewed chromosome breakage test results from 2010-2023 from a laboratory which receives samples from the majority of Australia’s six states and two territories, with an estimate coverage of 82.2% of Australia’s population. Tests were for the populations of Queensland (QLD), New South Wales (NSW), Victoria (VIC), Tasmania (Tas), the Australian Capital Territory (ACT) and the Norther Territory (NT), which accounts for 21,908.2 thousand residents of Australian’s population of 26638.5 thousand residents (source Australian Bureau of Statistics, June 30^th^, 2023). Testing data for 2023 was obtained from Western Australia with 23 tests performed and 1 being positive. Testing data for 2018-2024 was obtained from South Australia with 61 tests performed and 1 being positive.

### Screening for oral potentially malignant lesions

Non-invasive oral inspections and cytopathology for oral potentially malignant lesions were performed as described previously (Velleuer et al., 2020).

## Supplementary text and figures

### An estimation of the prevalence of FA in Australia

With a number of conservative assumptions we extrapolated the number of cases of FA in Australia to be approximately 189 or a frequency of 1 in 141,169 people (see Supplementary Text for calculations and assumptions). 58 abnormal (positive) tests were obtained over 14 years from the largest data set, which was from the Queensland laboratory. This works out to 4.14 cases of FA in Australia per year (58/14 = 4.14). Given that 82.2% of the population is estimated to be represented by this testing data, we need to perform an adjustment. We assume that the rate of FA in WA and SA is similar to the rate in the other states of Australia and would therefore need to apply a correction factor of 1.22 (100/82.2). Therefore, the adjusted estimate of FA cases per year is approximately 5.1 (4.14 x 1.22).

We estimated the total number of cases of FA in Australia by multiplying the adjusted estimate of FA cases per year by the median survival. While acknowledging the limitations of incomplete testing data and the limitations of using median survival age, which likely does not represent the mean due to an asymmetric survival distribution, the median provides a robust measure in this context. With a median survival of 37 years (95% CI: 34.7-43.1) for someone with FA (Altintas et al., 2022) we estimate that there could be approximately 189 cases of FA in Australia (5.1 x 37 = 188.7, 95% CI: 177.0-219.8), or 1 case per 141,169 people in Australia. This figure likely underestimates the number of cases because there were more inconclusive tests than abnormal tests in the dataset. Further, there may have been diagnostic tests for FA for residents of the six states or territories covered here which were sent to testing laboratories other than Queensland.

**Supplementary Figure 1.**
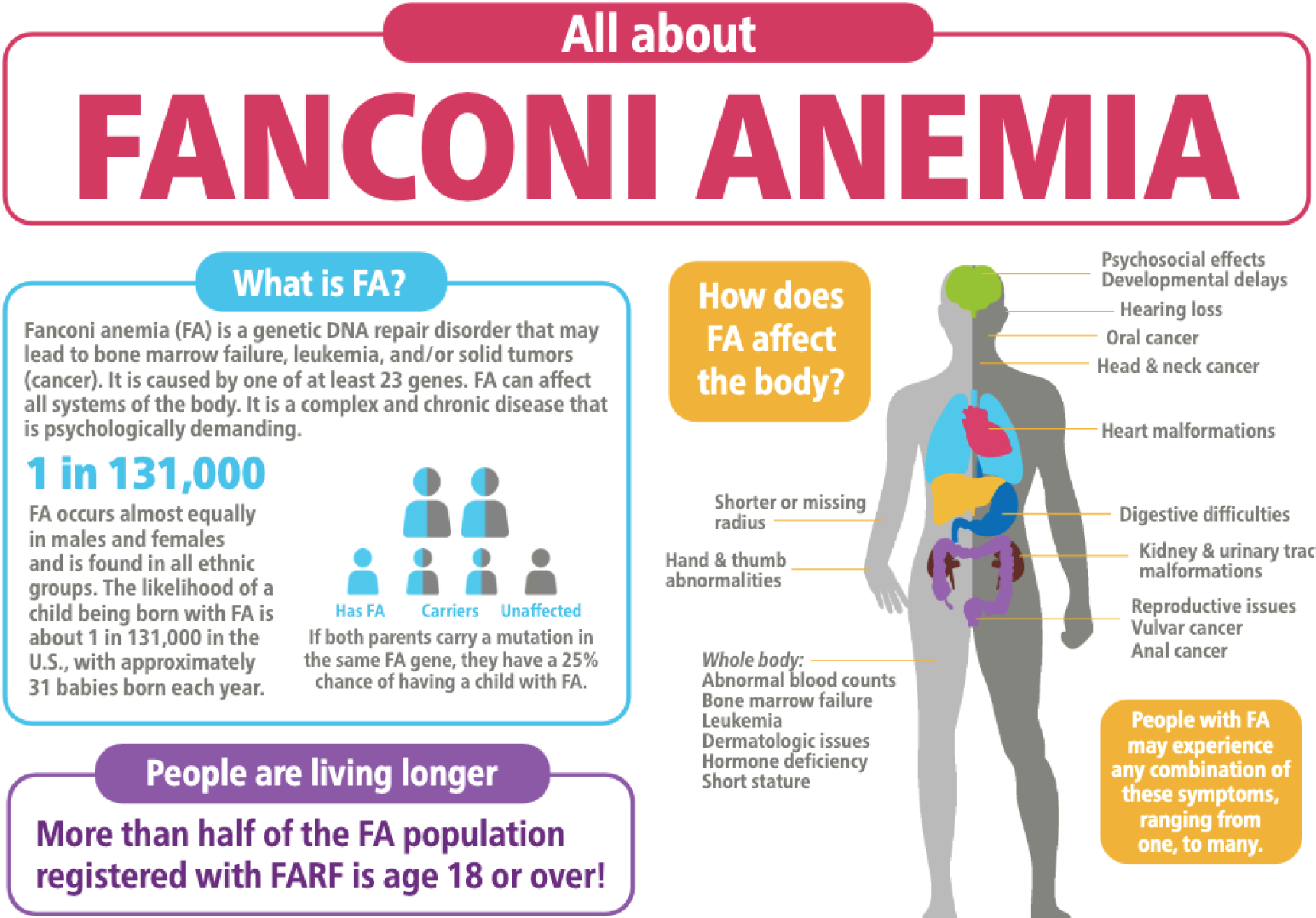
**Infographic: All About FA from the Fanconi Cancer Foundation**, which provides a practical graphical summary of potential issues that may be used to trigger suspicion of FA, which can be tested for with a chromosome breakage assay. Reproduced with permission. Source Fanconi.org.

